# Racial Disparities in the SOFA Score Among Patients Hospitalized with COVID-19

**DOI:** 10.1101/2021.03.31.21254735

**Authors:** Benjamin Tolchin, Carol Oladele, Deron Galusha, Nitu Kashyap, Mary Showstark, Jennifer Bonito, Michelle C. Salazar, Jennifer L. Herbst, Steve Martino, Nancy Kim, Katherine A. Nash, Max Jordan Nguemeni Tiako, Shireen Roy, Karen Jubanyik

**Author notes:** **Corresponding Author Contact Information:** (BT).

## Abstract

**Background:** Sequential Organ Failure Assessment (SOFA) score predicts probability of in-hospital mortality. Many crisis standards of care use SOFA score to allocate medical resources during the COVID-19 pandemic.

**Research Question:** Are SOFA scores disproportionately elevated among Non-Hispanic Black and Hispanic patients hospitalized with COVID-19, compared to Non-Hispanic White patients?

**Study Design and Methods:** Retrospective cohort study conducted in Yale New Haven Health System, including 5 hospitals with total of 2681 beds. Study population drawn from consecutive patients aged ≥18 admitted with COVID-19 from March 29^th^ to August 1^st^, 2020. Patients excluded from the analysis if not their first admission with COVID-19, if they did not have SOFA score recorded within 24 hours of admission, if race and ethnicity data were not Non-Hispanic Black, Non-Hispanic White, or Hispanic, or if they had other missing data. The primary outcomes was SOFA score, with peak score within 24 hours of admission dichotomized as <6 or ≥6.

**Results:** Of 2982 patients admitted with COVID-19, 2320 met inclusion criteria and were analyzed, of whom 1058 (45.6%) were Non-Hispanic White, 645 (27.8%) were Hispanic, and 617 (26.6%) were Non-Hispanic Black. Median age was 65.0 and 1226 (52.8%) were female. In univariate logistic screen and in full multivariate model, Non-Hispanic Black patients but not Hispanic patients had greater odds of an elevated SOFA score ≥6 when compared to Non-Hispanic White patients (OR 1.49, 95%CI 1.11-1.99).

**Interpretation:** Crisis standards of care utilizing the SOFA score to allocate medical resources would be more likely to deny these resources to Non-Hispanic Black patients.

## Introduction

Prior to the first wave of Coronavirus-2019 (COVID-19), models predicted that a pandemic respiratory virus might require ventilators, intensive care unit (ICU) beds, and other life-sustaining medical resources far in excess of available supplies. (1) On January 30^th^ 2020, the World Health Organization (WHO) declared a Public Health Emergency of International Concern which, in some countries, led to formal and informal restrictions on the allocation of critical medical resources on the basis of advanced age. (2, 3)

In response to early shortages and high rates of infection and mortality in Europe and the Northeastern United States, a number of healthcare systems and states in the US developed crisis standards of care (CSC): guidelines that advise hospitals and providers how to operate in a public health disaster, outside of their normal operating standards of care. CSC include triage protocols for the allocation of scarce life-sustaining medical resources. (4-8) The primary goal of published protocols is to establish a consistent system for allocating resources to save as many lives as possible during public health emergencies.

Publicly available triage protocols, prior to and during the pandemic, focus primarily on the Sequential Organ Failure Assessment (SOFA) score to assess patients’ likelihood of benefiting (surviving) as a result of receiving medical resources. (9) The SOFA score is a validated prognostic score ranging from 0-24, with points assigned for evidence of organ failure within 6 different organ systems, with higher scores correlating with a higher likelihood of in-hospital mortality. (10, 11) Originally developed and validated among septic patients in the medical ICU, the SOFA score has also been shown to predict mortality among patients with acute respiratory distress syndrome in the setting of COVID-19 infection. (12) Most disaster triage protocols prioritize patients who require medical resources but have lower SOFA scores to receive resources, on the grounds that such patients are more likely to benefit (survive).

In addition to threatening to overwhelm existing medical resources, the COVID-19 pandemic has also highlighted and exacerbated existing racial, ethnic, and socioeconomic health disparities. Marginalized populations, including racial and ethnic minorities and individuals of lower socioeconomic status, are more likely to become infected with COVID-19, more likely to be hospitalized, and more likely to die as a result. (13-15) Disparities in social determinants of health, including safe access to adequate nutritious food, exercise options, stable housing, and economic opportunities likely contribute to disparities in COVID-19 outcomes.

Marginalized populations are more likely to work in service-sector jobs that cannot be conducted remotely, are more likely to depend on public transportation, and are more likely to live in small and densely packed housing units and in group-living situations including homeless shelters, prisons, jails, and detention facilities. (16-19) They are less likely to have access to preventive healthcare and more likely to experience bias when they do access the healthcare system, resulting in higher rates of chronic comorbidities including diabetes, hypertension, and chronic pulmonary diseases. (20, 21) These pervasive inequities constitute a structure of systemic racism and contribute to higher rates of COVID-19 infection, more severe acute illness due to preexisting conditions, and higher mortality rates. (22, 23)

Given that marginalized populations are more likely to become sicker with COVID-19, utilization of CSC triage protocols, which rely on the SOFA score, have the potential to disproportionately deny medical resources to racial and ethnic minorities. (24, 25) The potential for triage protocols to exacerbate racial and ethnic health disparities has been documented in patient cohorts with sepsis and acute respiratory distress syndrome (ARDS) but has not previously been examined in patients with COVID-19. (26) There is therefore a lack of evidence as to whether there are disparities by race and ethnicity in SOFA scores amongst patients admitted with COVID-19. We conducted a retrospective cohort study to determine whether SOFA scores are disproportionately elevated among members of racial and ethnic minorities, and specifically Non-Hispanic Black and Hispanic patients, in comparison to Non-Hispanic White patients with COVID-19. The existence of such a disparity would raise significant concerns about the use of triage protocols relying on SOFA scores and the potential for exacerbating racial and ethnic health inequities during future waves of the COVID-19 pandemic and other public health emergencies.

## Methods

### Study design and data source

We conducted a retrospective cohort study of patients with COVID-19 within the Yale-New Haven Health System (YNHH) from March 29th, 2020 to August 1, 2020. YNHH includes 5 hospitals and a large physician practice base, serving racially, ethnically, and socioeconomically diverse communities across Connecticut and Rhode Island. The hospitals range from primary community hospitals to a tertiary academic medical center, with a total of 2,681 beds. Data from the YNHH electronic medical record (EMR, Epic Systems Corporation, Verona, WI) database was used for analyses. The study was approved by the Yale University Human Subjects Committee (study number 2000028081).

### Participants

We included EMR data for all patients age ≥18 with COVID-19 admitted to YNHH hospitals during the study period. Patients were considered positive for COVID-19 if they had a positive PCR test or clinical markers including fever, cough and chest radiographs considered to be consistent with COVID-19 infection in the setting of the first wave of the pandemic in the northeastern United States, and designated as COVID-19 positive by an attending physician. Patients <18 years of age were excluded as the SOFA score is not validated in pediatric patients. Patients were excluded from the analysis if they did not have a SOFA score recorded within 24 hours of admission, if it was not their first admission with COVID-19, or if their race and ethnicity data were not Non-Hispanic Black, Non-Hispanic White, or Hispanic (Fig 1). Because prior COVID-19 studies show that Black and Hispanic patients experience higher rates of critical illness and mortality, (13-15) we hypothesized that Non-Hispanic Black and Hispanic patients will be more likely to have elevated SOFA scores within 24 hours of admission compared to Non-Hispanic White patients.

**Fig 1.**
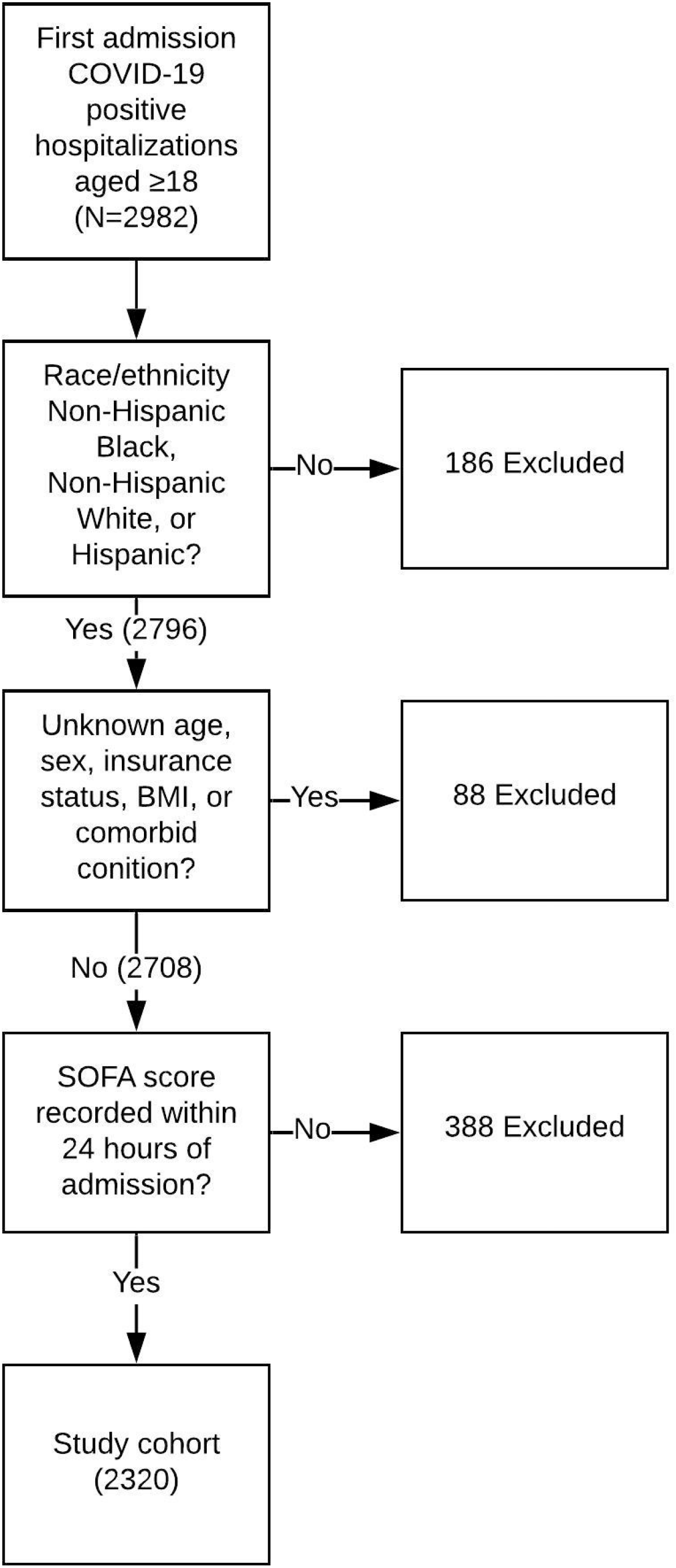
Construction of Study Cohort. Abbreviations: BMI: Body Mass Index; COVID-19: Coronavirus Disease 2019; SOFA: Sequential Organ Failure Assessment.

### Predictor variables

Data extracted from the EMR included sociodemographic and clinical variables. Our main predictor variables were age, sex, race, ethnicity, and insurance status. These variables are recorded by admitting clerks at YNHH hospitals. Other variables included clinical characteristics like body mass index (BMI) and comorbid conditions known to be associated with mortality in the setting of COVID-19. (15) Smoking status was not included in the analysis, because in our clinical experience there is a significant desirability bias, leading patients to report themselves to clinicians as non-smokers or former smokers rather than current smokers. (27)

### Outcome variable

The main outcome, SOFA score, was continuously and automatically calculated for all admitted patients and recorded every 4 hours within the YNHH EMR. SOFA score was determined by an automated algorithm within the EMR system, assigning 0-4 points for each of 6 organ systems (neurologic, pulmonary, cardiovascular, renal, hepatic, hematologic), based on laboratory, respiratory and nursing flowsheet data in the EMR, following previously specified and validated rules.(10) The total SOFA score ranges from 0-24, with higher scores indicating a higher likelihood of in-hospital mortality. A binary SOFA variable (peak score within 24 hours <6, ≥6) was created to examine variation in illness severity by patient sociodemographic characteristics. We focused on this dichotomous outcome because published triage protocols categorize patients with a SOFA score <6 as being in the most prioritized group, most likely to receive scarce medical resources in a disaster situation, whereas patients with SOFA score ≥6 are deprioritized, resulting in lower likelihoods of receiving scarce medical resources.(4, 8) We focused on peak SOFA score within the first 24 hours because in a public health emergency in which life-sustaining medical resources are fully occupied, it is initial SOFA scores that will determine whether a newly admitted critically ill patient receives scarce resources.

### Statistical analysis

We used Analysis of Variance (ANOVA) tests to examine mean differences in peak 24-hour SOFA score by sociodemographic and clinical characteristics. Chi-square tests were used to examine differences in the proportion of COVID positive patients with SOFA score ≥6 and <6 by patient characteristics. Finally, we conducted logistic regression analyses to assess racial differences in SOFA score adjusting for sociodemographic and clinical covariates. We considered candidate covariates based on clinical experience and emerging evidence regarding associations with clinical outcomes in COVID-19. The final multivariate model was then refined through the exclusion of collinear covariates. We conducted a univariate screen followed by a multivariate regression adjusting for all sociodemographic and clinical covariates listed in Table 3. Race-stratified models were also constructed to assess whether factors associated with SOFA score varied according to race and ethnicity.

**Table 3:** Univariate and multivariate regression model results for factors associated with SOFA score within 24 hours ≥ 6

| Characteristic | Model 1- Combined Univariate (unadjusted) |  |  |  | Model 1- Combined Multivariate |  |  |  |
| --- | --- | --- | --- | --- | --- | --- | --- | --- |
|  | OR | 95% CI |  | p-value | OR | 95% CI |  | p-value |
| Race/Ethnicity |  |  |  |  |  |  |  |  |
| Hispanic | 0.78 | 0.58 | 1.06 | 0.1077 | 0.87 | 0.62 | 1.24 | 0.4501 |
| Black Non-Hispanic | 1.58 | 1.21 | 2.06 | 0.0008* | 1.49 | 1.11 | 1.99 | 0.0075* |
| White Non-Hispanic | reference |  |  |  | Reference |  |  |  |
| Age |  |  |  |  |  |  |  |  |
| 18-34 | reference |  |  |  | Reference |  |  |  |
| 35-64 | 2.55 | 1.44 | 4.52 | 0.0013* | 1.95 | 1.07 | 3.54 | 0.0288* |
| >=65 | 2.84 | 1.62 | 4.98 | 0.0003* | 2.57 | 1.32 | 4.98 | 0.0052* |
| Sex |  |  |  |  |  |  |  |  |
| Men | 1.83 | 1.45 | 2.32 | <.0001* | 1.94 | 1.51 | 2.50 | <.0001* |
| Women | Reference |  |  |  | Reference |  |  |  |
| Insurance status |  |  |  |  |  |  |  |  |
| Private | Reference |  |  |  | Reference |  |  |  |
| Medicare | 1.53 | 1.09 | 2.14 | 0.0135* | 1.07 | 0.70 | 1.63 | 0.757 |
| Medicaid | 1.40 | 0.94 | 2.07 | 0.0955 | 1.35 | 0.89 | 2.04 | 0.1536 |
| Uninsured | 1.01 | 0.58 | 1.76 | 0.9678 | 1.19 | 0.66 | 2.13 | 0.566 |
| BMI |  |  |  |  |  |  |  |  |
| <25 | Reference |  |  |  | Reference |  |  |  |
| 25 - 29.9 | 1.07 | 0.79 | 1.45 | 0.6708 | 1.22 | 0.88 | 1.68 | 0.2317 |
| 30-34.9 | 1.08 | 0.77 | 1.52 | 0.6628 | 1.30 | 0.91 | 1.87 | 0.1561 |
| 35+ | 1.17 | 0.85 | 1.62 | 0.3372 | 1.52 | 1.07 | 2.18 | 0.0207* |
| Comorbid conditions |  |  |  |  |  |  |  |  |
| Chronic pulmonary disease | 1.27 | 0.99 | 1.62 | 0.0603 | 1.07 | 0.81 | 1.40 | 0.6453 |
| CHF | 1.86 | 1.45 | 2.38 | <.0001* | 1.19 | 0.86 | 1.63 | 0.288 |
| Diabetes | 1.65 | 1.31 | 2.08 | <.0001* | 1.07 | 0.82 | 1.41 | 0.6082 |
| CAD | 1.89 | 1.48 | 2.42 | <.0001* | 1.18 | 0.86 | 1.61 | 0.3151 |
| Hypertension | 1.67 | 1.27 | 2.19 | 0.0003* | 0.95 | 0.67 | 1.34 | 0.7594 |
| Advance renal disease | 3.15 | 2.29 | 4.34 | <.0001* | 2.35 | 1.62 | 3.40 | <.0001* |
| Advance liver disease | 2.96 | 1.58 | 5.54 | 0.0007* | 2.51 | 1.29 | 4.89 | 0.0068* |

Abbreviations: \*: p-value < 0.05; BMI: Body Mass Index; CAD: Coronary Artery Disease; CHF: Congestive Heart Failure; CI: Confidence Interval; OR: Odds Ratio; SOFA: Sequential Organ Failure Assessment.

## Results

From March 29^th^ to August 1^st^, there were 3362 admissions of COVID-19-positive patients aged ≥18 to YNHH hospitals. Of these, 2982 were first admissions (Fig. 1) and 2796 were Non-Hispanic Black, Non-Hispanic White, or Hispanic. Of these, 88 had missing baseline demographics or clinical data, and 388 had missing SOFA scores, and were excluded. Two thousand three hundred and twenty patients had complete race/ethnicity and baseline characteristics, were either Hispanic, Non-Hispanic Black, or Non-Hispanic White, and were included in the analysis. There were no statistically significant differences in demographic or clinical characteristics between patients with and without SOFA scores.

Within the study cohort of 2320, 1058 (45.6%) were Non-Hispanic White, 645 (27.8%) were Hispanic, and 617 (26.6%) were Non-Hispanic Black (Table 1). The median age was 65.0, and 1226 (52.8%) were female. Six-hundred and fifty-nine (28.4%) had Medicaid or no insurance. Nine-hundred and sixty-nine (41.7%) were obese. A total of 1829 (78.8%) had one or more comorbid conditions thought to increase risk of mortality in the setting of COVID-19. Patients with peak SOFA scores ≥6 within the first 24 hours were disproportionately common among Non-Hispanic Black patients, older patients, males, and patients with Congestive Heart Failure (CHF), diabetes, coronary artery disease (CAD), hypertension, advanced renal disease, and advanced liver disease. Baseline characteristics broken down by race/ethnicity are available (S1 Table).

**Table 1:**
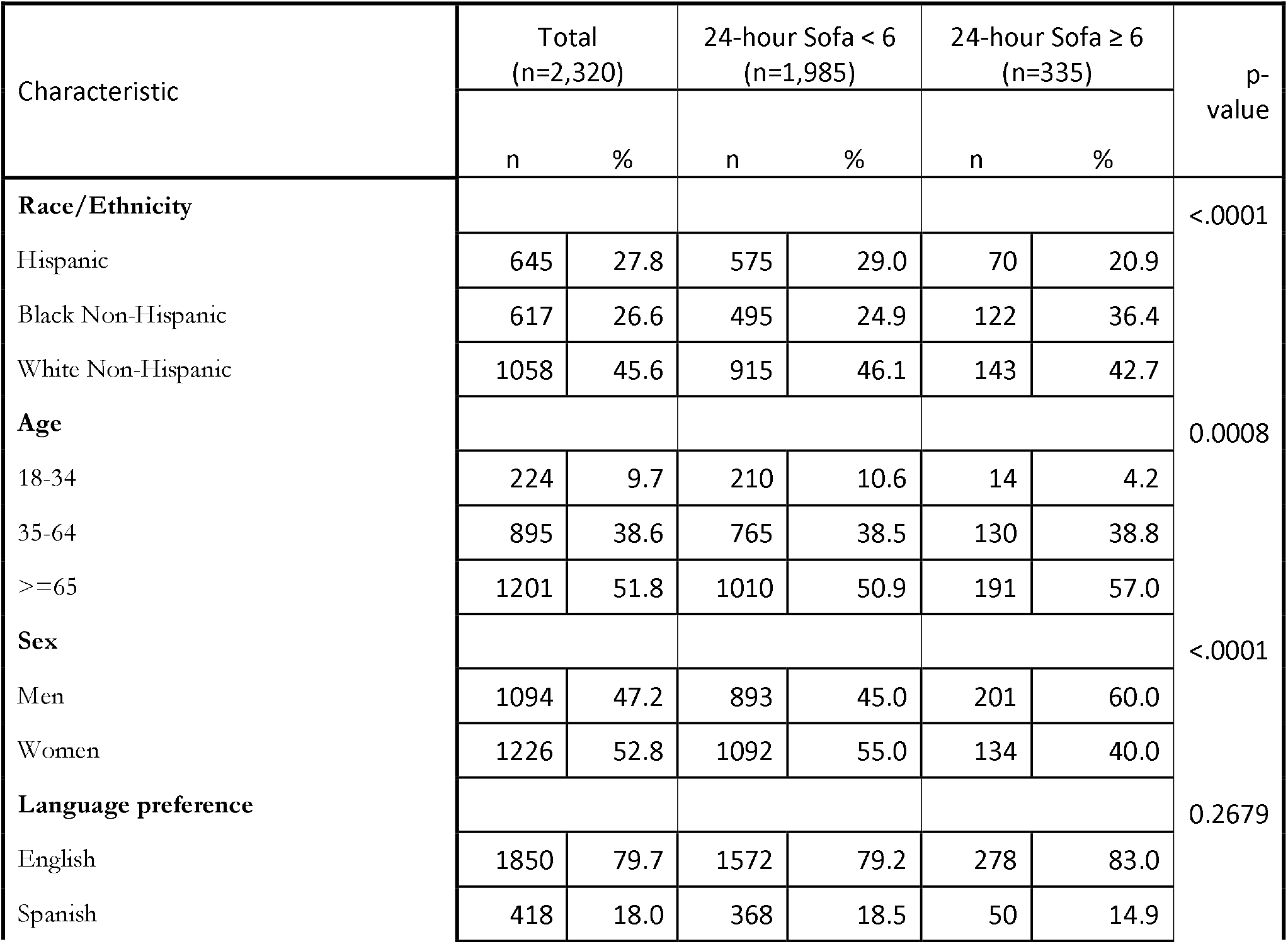
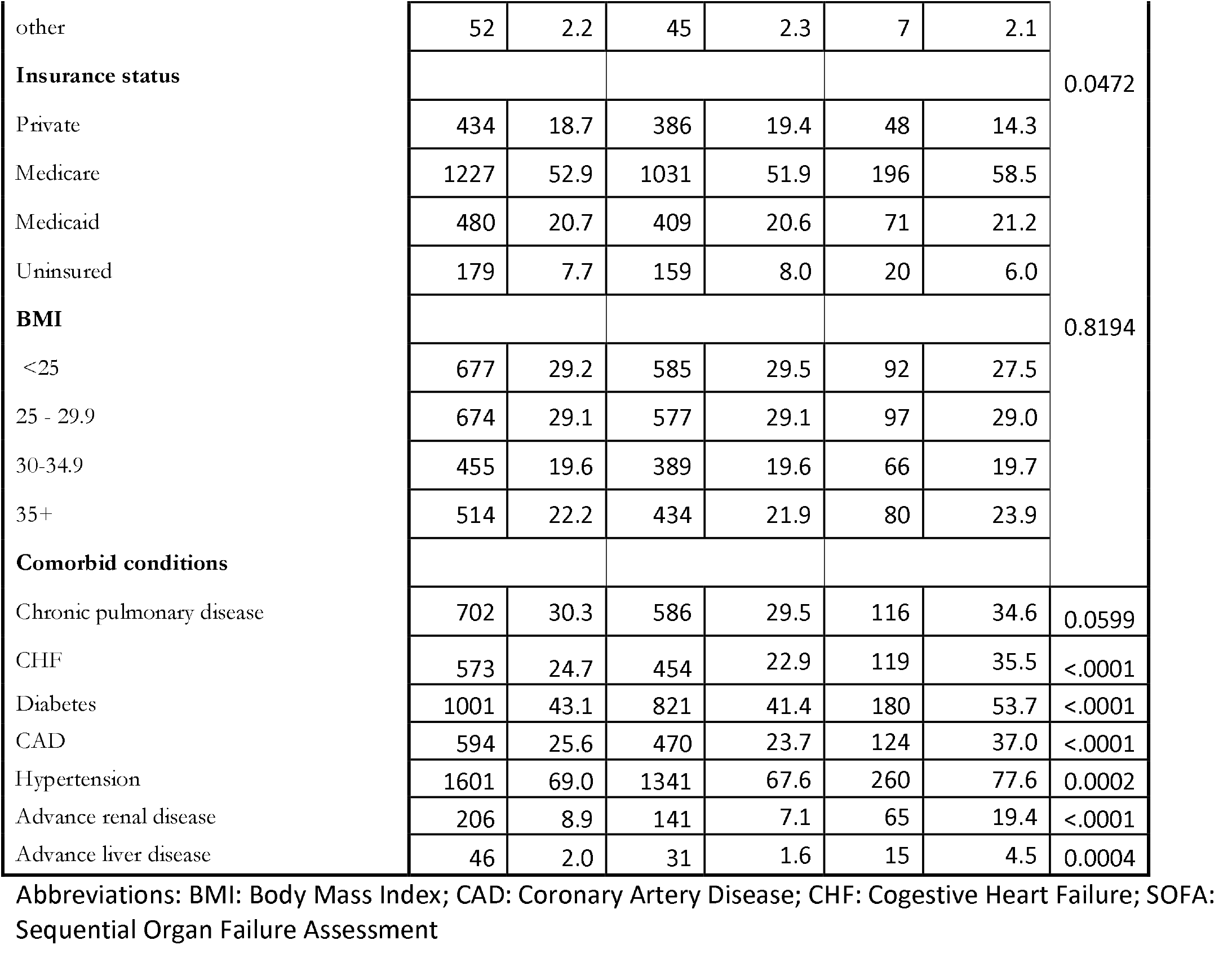
Characteristics of COVID+ patients with SOFA within 24 hours of admission; SOFA <6 and ≥6

Mean peak SOFA score within the first 24 hours was (2.4±3.0) overall, ranging from 0 to 18 (Table 2). Mean SOFA score was significantly elevated among Non-Hispanic Black patients (3.0±3.1), but not among Hispanic patients (2.2±3.1) in comparison to Non-Hispanic White patients (2.5±2.8). SOFA score was also significantly elevated among patients aged 35-64 (2.5±3.0) and ≥65 (2.8±3.0) in comparison to those aged 18-34 (1.3±2.3), among Men (3.0±3.2) in comparison to Women (2.2±2.6), and among those with Medicare insurance (2.9±3.0) but not Medicaid (2.3±3.0), or no insurance (2.0±3.1) compared to those with private insurance (2.0±2.8). The SOFA score was also significantly elevated among those with comorbid conditions including CHF (3.4±3.2) compared to those without (2.3±2.8), diabetes (3.0±3.1) compared to those without (2.2±2.8), CAD (3.3±3.2) compared to those without (2.3±2.8), hypertension (2.8±3.0) compared to those without (1.9±2.7), advanced renal disease (4.7±3.1) compared to those without (2.3±2.9), and advanced liver disease (4.8±3.9) compared to those without (2.5±2.9).

**Table 2:**
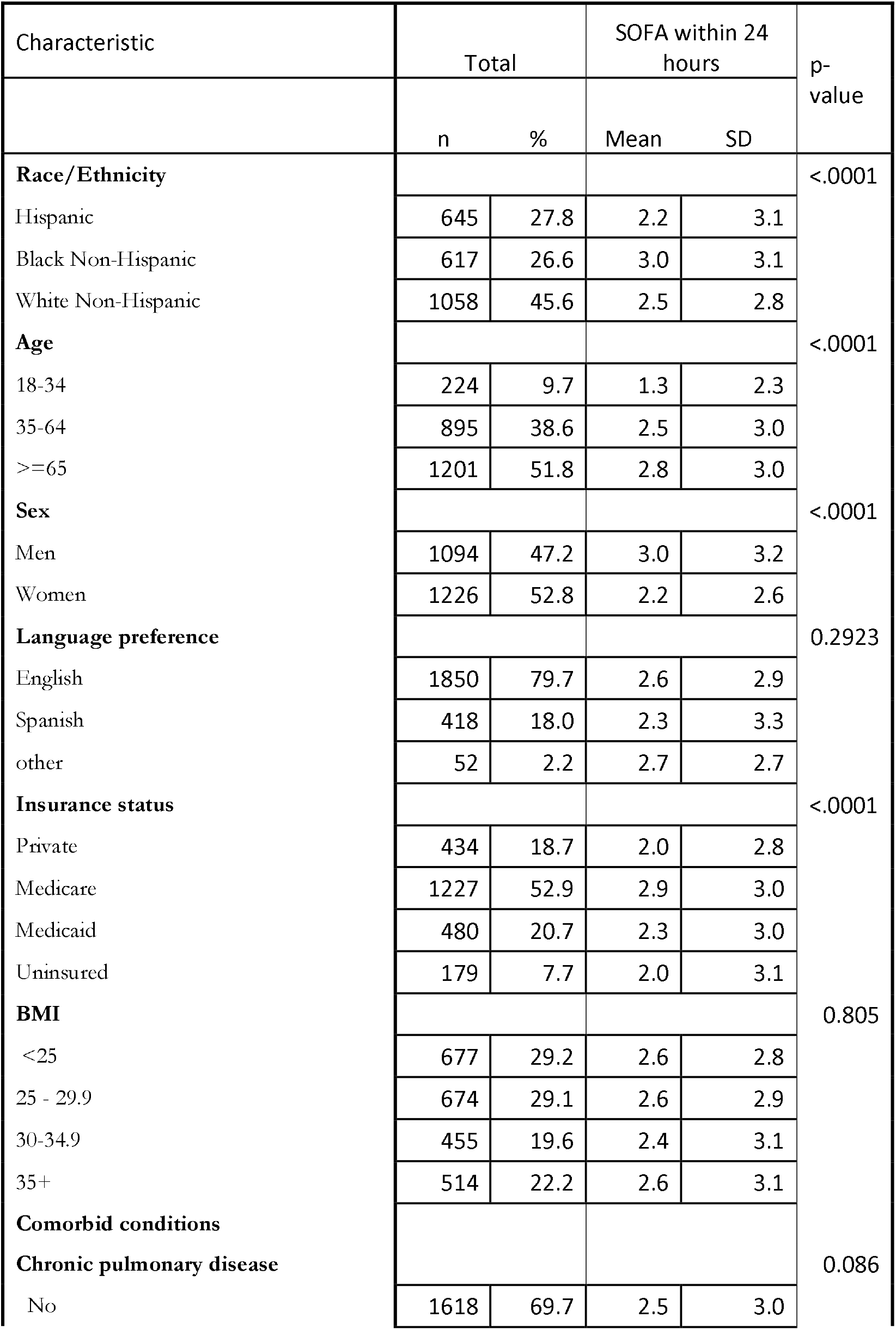
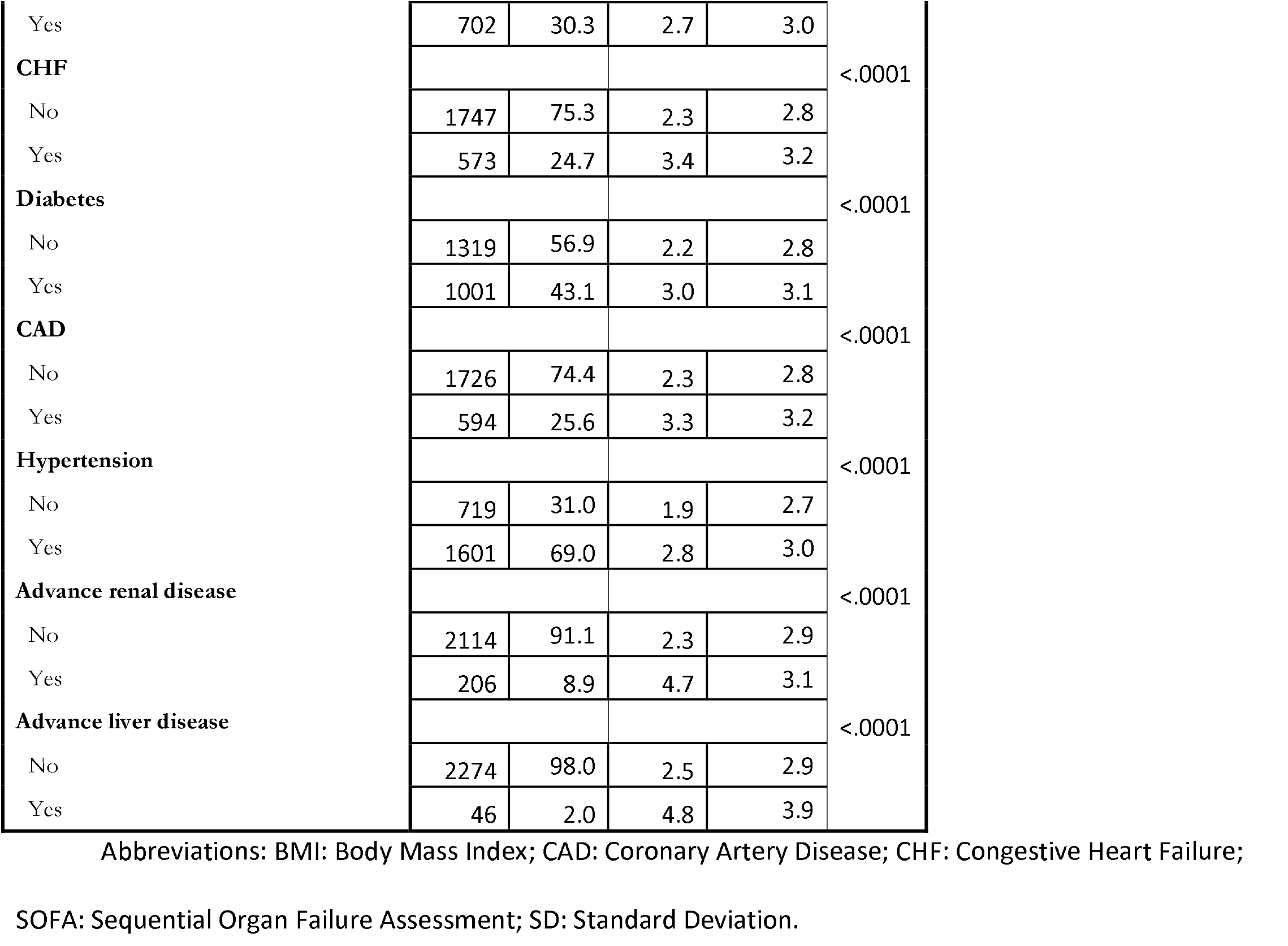
Mean SOFA within 24 hours of admission.

In a univariate logistic screen and in a full multivariate model (Table 3), Non-Hispanic Black patients had greater odds of an elevated SOFA score ≥6 when compared to Non-Hispanic White patients (OR 1.49, 95%CI 1.11-1.99). In contrast, Hispanic patients did not have increased odds of an elevated SOFA score. Advanced age was also associated with increased odds of elevated SOFA score (OR 1.95, 95%CI 1.07-3.54 for age 35-64, OR 2.57, 95%CI 1.32-4.98 for age ≥65), as was male sex (OR 1.94, 95%CI 1.51-2.50), body-mass index ≥35 (OR 1.52, 95%CI 1.07-2.18), advanced renal disease (OR 2.35, 95%CI 1.62-3.40), and advanced liver disease (OR 2.51, 95%CI 1.29-4.89). Medicare was associated with increased odds of elevated SOFA score, but dropped out in the multivariate model, when other variables such as age were included. Race stratified models were also constructed but did not identify new covariates associated with elevated SOFA scores in both univariate screen and multivariate logistic analysis. We reran the analysis looking at peak 48 hour SOFA score with unchanged results.

## Discussion

In our cohort of COVID-19 positive patients admitted to YNHH hospitals, Non-Hispanic Black race/ethnicity, male sex, advanced age, stage II or greater obesity, advanced renal disease, and advanced liver disease were all independently associated with significantly higher odds of elevated peak SOFA score ≥ 6 during the first 24-hours of admission. Hispanic ethnicity was not associated with increased risk of elevated SOFA score. Medicaid and Medicare insurance types were not independently associated with increased odds of elevated SOFA score.

These findings are consistent with prior studies showing that Black race, older age, obesity, and chronic medical comorbidities are associated with increased rates of mortality in COVID-19. (15) These findings ar e also consistent with prior findings that SOFA overestimates mortality among Black patients and underestimates mortality among White patients with sepsis and ARDS prior to the COVID-19 pandemic. (26) The racial disparities in SOFA scores we found among patients with COVID might be due to this systemic overestimation of mortality among Black persons and underestimation of mortality among White persons. Alternatively, Black persons with COVID-19 might have higher SOFA scores in the hospital because COVID-19 affects them more severely, for example because they are subjected to higher levels of discrimination and stress or because they have less access to long-term preventive care. (20, 21, 28) Finally, Black patients might have higher SOFA scores at the time of admission because they present or are admitted to hospitals only when they are sicker.(29, 30) This could be because of current or prior discrimination within the healthcare system that might discourage patients from seeking medical attention with mild or moderate symptoms. (31) Our data does not directly explain the cause of elevated SOFA scores among Black patients with COVID-19.

It is notable that patients with Medicaid or no insurance did not have elevated SOFA scores in comparison to patients with private insurance. This might suggest that insurance status plays a relatively limited role in elevated SOFA scores during the first 24 hours of hospital admission. If so, other factors, such as housing density, public transportation utilization, employment in the service sector, telecommuting opportunities, racial discrimination, or distrust of the healthcare system, might account for elevated SOFA scores among Non-Hispanic Black patients.

Because published triage protocols utilize the SOFA score to allocate scarce medical resources, and prioritize patients with SOFA score <6 over other patients, such protocols − if implemented − would be more likely to triage Non-Hispanic Black people to not receive scarce resources such as ventilators and ICU beds during future waves of the COVID-19 pandemic. As part of a system that predictably leads to racial disparities in health outcomes, triage protocols have the potential to become a component of systemic racism.

Given these findings and the possibility that crisis standards of care may be implemented during the COVID-19 pandemic, it is important to prospectively consider and implement measures to reduce systemic racism, protect marginalized populations, and promote racial and ethnic equity. The ideal would be to minimize or prevent entirely the need for triage, particularly among marginalized populations. This might be achieved in the short term through public health education, distribution of personal protective equipment, stockpiling of critical medical resources, targeted COVID-19 testing, contact tracing, social distancing, and even lockdowns coupled with financial support. The manifest injustice of the systemic racism and health inequities that COVID-19 has highlighted should also motivate long-term efforts to achieve more equitable health outcomes in the United States. These might include universal health insurance, a more redistributive system of taxation, housing support, elimination of food deserts and neighborhood segregation, anti-racism trainings for clinicians, and recruitment of marginalized populations into the medical workforce.

It is also possible to make crisis standards of care and triage protocols themselves more equitable. The development, revision, and oversight of these protocols might be made more open and transparent to patients, community members, and to the general public. Healthcare systems and states might recruit triage advisory and oversight committees that specifically include robust representation from ethnic and racial minorities, as well as individuals with disabilities and other marginalized populations. (8) Committees might specifically recruit advocacy organizations, faith leaders, institutional diversity officers, and other community leaders to ensure adequate representation of community concerns. The triage teams that implement protocols in hospitals might also be mandated to include representation of diverse perspectives.

In addition, the SOFA score might be supplemented to achieve more equitable outcomes. Prioritarian triage protocols might still use mortality probability scoring, such as the SOFA score, but might give marginalized populations a bonus or prioritization in these assessments. For example, patients might have their priority score improved slightly on the basis of their home address, using the Area Deprivation Index. (32) Potential comparative advantages and disadvantages of alternative triage systems are reviewed elsewhere. (33)

Our study is limited in that it was conducted within a single healthcare system in the Northeastern United States. Our healthcare system experienced a surge of COVID patients relatively early in the pandemic, with a peak on April 22, 2020 followed by relatively lower numbers, and medical care for COVID-19 has evolved over the course of the pandemic. While YNHH serves significant Hispanic and Non-Hispanic Black patient populations, it serves relatively smaller numbers of Asian, Native American, Pacific Islander, and other patient populations, and these small samples statistically forbade inclusion in the analysis. The disparities that this study documents may not be generalizable to other regions with different racial and ethnic demographics within the United States or globally. Our study is also limited by the data available within the clinical EMR. For example, racial and ethnic data is generally documented by unit clerks based on their observation of patients rather than on patient’s self-identification. Prior studies have shown that “socially assigned” race does associate closely with health outcomes. (34) Another limitation is that we did not investigate potential disparities in SOFA scores in other marginalized populations. Future research is needed to examine the effects of disability, psychiatric comorbidities, substance use disorders, unstable housing, or incarceration on SOFA scores.

In conclusion, Non-Hispanic Black patients admitted to hospitals with COVID-19 had increased odds of an elevated SOFA score ≥6 within the first 24-hours of admission. Therefore, published triage protocols utilizing the SOFA score to allocate scarce medical resources would be more likely to deny Non-Hispanic Black patients scarce medical resources such as ventilators and ICU beds if implemented during the COVID-19 pandemic. Governments and healthcare systems should prospectively consider and implement measures to reduce systemic racism, protect marginalized populations, and promote racial and ethnic equity the pandemic.

## Data Availability

Data cannot be shared publicly because this is human subjects research involving patient health information that is not directly identifying but does include indirect identifiers (such as sex, ethnicity, age category, insurance status, hospitalization dates, etc.). These indirect identifiers in combination may become identifying. Data are available from the Yale University Human Subjects Committee (203-785-4688) for researchers who meet the criteria for access to confidential data.

## Acknowledgements

The authors wish to acknowledge the support of the Center for Medical Informatics and the Equity Research and Innovation Center at Yale School of Medicine. In particular we are indebted to Indira Flores, Rebecca Vergara Greeno, and Olamide Olawoyin for assisting in literature review and project planning.

---

**S1 Table. Characteristics of COVID+ patients by Race/Ethnicity**. Abbreviations: BMI: Body Mass Index; CAD: Coronary Artery Disease; CHF: Cogestive Heart Failure; SOFA: Sequential Organ Failure Assessment

