## Supplemental Table 1 for "Racial Disparities in the SOFA Score Among Patients Hospitalized with COVID-19"

S1 Table: Characteristics of COVID+ patients by Race/Ethnicity

| Characteristic | Total (n=2,320) | | Hispanic (n=645) | | Non-Hispanic Black  (n=617) | | Non-Hispanic White  (n=1,058) | | p-value |
| --- | --- | --- | --- | --- | --- | --- | --- | --- | --- |
|  | n | % | n | % | n | % | N | % |  |
| **Age** |  |  |  |  |  |  |  |  | <.0001 |
| 18-34 | 224 | 9.7 | 121 | 18.8 | 58 | 9.4 | 45 | 4.3 |  |
| 35-64 | 895 | 38.6 | 344 | 53.3 | 284 | 46.0 | 267 | 25.2 |  |
| >=65 | 1201 | 51.8 | 180 | 27.9 | 275 | 44.6 | 746 | 70.5 |  |
| **Sex** |  |  |  |  |  |  |  |  | 0.0002 |
| Men | 1094 | 47.2 | 348 | 54.0 | 268 | 43.4 | 478 | 45.2 |  |
| Women | 1226 | 52.8 | 297 | 46.0 | 349 | 56.6 | 580 | 54.8 |  |
| **Language preference** |  |  |  |  |  |  |  |  | <.0001 |
| English | 1850 | 79.7 | 226 | 35.0 | 597 | 96.8 | 1027 | 97.1 |  |
| Spanish | 418 | 18.0 | 415 | 64.3 | 0 | 0.0 | 3 | 0.3 |  |
| other | 52 | 2.2 | 4 | 0.6 | 20 | 3.2 | 28 | 2.6 |  |
| **Insurance status** |  |  |  |  |  |  |  |  | <.0001 |
| Private | 434 | 18.7 | 116 | 18.0 | 134 | 21.7 | 184 | 17.4 |  |
| Medicare | 1227 | 52.9 | 166 | 25.7 | 296 | 48.0 | 765 | 72.3 |  |
| Medicaid | 480 | 20.7 | 224 | 34.7 | 158 | 25.6 | 98 | 9.3 |  |
| Uninsured | 179 | 7.7 | 139 | 21.6 | 29 | 4.7 | 11 | 1.0 |  |
| **BMI** |  |  |  |  |  |  |  |  | <.0001 |
| <25 | 677 | 29.2 | 153 | 23.7 | 151 | 24.5 | 373 | 35.3 |  |
| 25 - 29.9 | 674 | 29.1 | 211 | 32.7 | 148 | 24.0 | 315 | 29.8 |  |
| 30-34.9 | 455 | 19.6 | 152 | 23.6 | 129 | 20.9 | 174 | 16.4 |  |
| 35+ | 514 | 22.2 | 129 | 20.0 | 189 | 30.6 | 196 | 18.5 |  |
| **Comorbid conditions** |  |  |  |  |  |  |  |  |  |
| Chronic pulmonary disease | 702 | 30.3 | 137 | 21.2 | 222 | 36.0 | 343 | 32.4 | <.0001 |
| CHF | 573 | 24.7 | 68 | 10.5 | 177 | 28.7 | 328 | 31.0 | <.0001 |
| Diabetes | 1001 | 43.1 | 230 | 35.7 | 354 | 57.4 | 417 | 39.4 | <.0001 |
| CAD | 594 | 25.6 | 81 | 12.6 | 171 | 27.7 | 342 | 32.3 | <.0001 |
| Hypertension | 1601 | 69.0 | 294 | 45.6 | 500 | 81.0 | 807 | 76.3 | <.0001 |
| Advance renal disease | 206 | 8.9 | 27 | 4.2 | 100 | 16.2 | 79 | 7.5 | <.0001 |
| Advance liver disease | 46 | 2.0 | 15 | 2.3 | 14 | 2.3 | 17 | 1.6 | 0.4917 |
| Charlson comorbidity index - mean, sd | 1.8 | 2.2 | 1.2 | 2.0 | 2.0 | 2.4 | 2.0 | 2.3 | <.0001 |

Abbreviations: BMI: Body Mass Index; CAD: Coronary Artery Disease; CHF: Cogestive Heart Failure; SOFA: Sequential Organ Failure Assessment
